# From Peak to Plunge: A Multi-Database Analysis of State Level Disparities in Hydromorphone use in the U.S. (2010-2023)

**DOI:** 10.1101/2025.07.24.25332070

**Authors:** Krisha S. Patel, Kenneth L. McCall, Brian J. Piper

## Abstract

**Background:** Hydromorphone is a semi-synthetic opioid agonist and a hydro-genated ketone of morphine. This study observed hydromorphone use in the United States (US) using three databases. Methods: The distribution of hydromorphone in the US (in grams) was provided by US Drug Enforcement Administration’s Automated Reports and Consolidated Orders System (ARCOS) by state, zip code, and by business types (pharmacies, hospitals, providers, etc.). Hydromorphone prescriptions claims were also examined using the Medicaid and Medicare Part D programs from 2010 to 2023. Results: Hydromorphone increased by +30.6% by 2013, followed by a decrease of −55.9% by 2023 in ARCOS. Medicaid hydromorphone prescriptions increased +39.6% by 2015 and de-creased −48.9% by 2023. Medicare Part D hydromorphone claims increased +8.5% by 2015 and decreased −31.9% by 2023. There were also pronounced regional disparities in hydro-morphone use identified in ARCOS (158.7 fold), Medicaid (17.5 fold), and Medicare Part D (13.7 fold). Conclusions: Hydromorphone use in the US has decreased substantially from 2010 to 2023. Additionally, these findings highlight regional disparities in hydro-morphone use, which may inform targeted opioid stewardship initiatives and guide policymakers to ensure safe and equitable opioid prescribing practices.

## 1. Introduction

Hydromorphone is a potent opioid approved for managing moderate-to-severe acute and severe chronic pain in patients [1], Table 1. Hydromorphone has a 5–10-fold greater analgesic effect than morphine and crosses the blood-brain barrier more easily [2]. Hydro-morphone is available orally in powder, solution, immediate-release tablet, modified-release, and parenterally by intravenous, intramuscular, and subcutaneous routes. It is absorbed in the upper small intestine, is extensively metabolized by the liver, and has a variety of renally excreted, water-soluble metabolites. Like morphine, it is a μ opioid receptor agonist and binds to a lesser degree to the delta receptors. Because it is more fat soluble than morphine, its onset of action is correspondingly faster than that of morphine but is slower than highly lipid soluble drugs such as fentanyl [3].

**Table 1.** Pharmacology properties for hydromorphone.

| Properties | Data | Source |
| --- | --- | --- |
| FDA approved for age | Severe pain (adults) | [11] |
| Pharmacodynamics | Opioid ($\mu$ agonist) | [12] |
| Formulations | immediate release and extended release tablets and capsules, oral solution, IV | [13] |
| US Schedule (UK Class) | II | [13] |
| Dose (mg/day) | 2-8<sup>I</sup>, 8-32<sup>E</sup> | [13] |
| Half-life (hours) | 8-15 | [12] |
| Streetrx.com cost/year (2023) | \$15,534.4<sup>I</sup>, \$62,137.6<sup>E</sup> | [6] |
| I: immediate release |  |  |
| E: extended release |  |  |

Hydromorphone is prescribed only when initial treatments have proven ineffective, primarily due to the drug’s elevated potency, potential for abuse, and risk of overdose [4]. It is a controlled (schedule II) substance. Acute overdose of hydromorphone can produce severe respiratory depression, somnolence progressing to stupor or coma, skeletal muscle flaccidity, cold and clammy skin, constricted pupils, reduction in blood pressure and heart rate, and death. [5] The Researched Abuse, Diversion and Addiction-Related Surveillance (RADARS) System StreetRx Program is based on crowdsourcing street prices of drugs. Site visitors spontaneously and anonymously submit the street prices they paid, or heard were paid, for diverted prescription drugs. As of 2023, based on the average prices mentioned on streetrx, (corrected by the annual rate of inflation), a one mg immediate release tablet of hydromorphone has a black market value of around 15,000 dollars per year, whereas a one mg extended-release tablet values around 62,000 dollars per year [6].

Past research has identified pronounced geographical disparities in several opioids including morphine, oxycodone, and codeine [7], but hydromorphone has not yet been studied. The purpose of this study was to identify temporal and state-level disparities in hydromorphone use in the US from 2010 to 2023. Three databases were used for this investigation: the Drug Enforcement Administration’s Automated Reports and Consolidated Orders System (ARCOS) [8], Medicaid [9], and Medicare Part D [10]. This study also examined correlations between these three datasets to better understand trends and relationships in hydromorphone distribution and prescriptions.

## 2. Materials and Methods

### Data sources

The US Drug Enforcement Administration’s Automated Reports and Consolidated Orders System (ARCOS) provided the distribution of hydromorphone (grams) nationally, by state, zip code, and by business types (pharmacies, hospitals, providers, etc.) [8]. The US Census Bureau provided the adults per state for 2013 and 2023 to correct the distribution for population [14].

Medicaid.gov provided the number of prescriptions (brand and generic) per state in the US from 2013-2023 [9]. Data.Medicaid.gov provided the number of Medicaid enrollees by state for December of 2015 and 2023 [15].

The US Centers for Medicare and Medicaid Services (CMS) provided the number of Medicare Part D claims nationally and per state for brand and generic formulations for 2013 to 2023 [10]. CMS also provided the number of Medicare enrollees by state, in December of 2015 and 2023 to correct the prescriptions for the enrollees [16]. The US Physician Workforce Data Dashboard provided the number of physicians by state in the US in 2023 [17]. The US Census Bureau provided information on the demographics of the population by state distributed by race (Whites/non-white) in the US [18].

### Statistical analysis

We used Datawrapper to create heatmaps [19], GraphPad Prism (Version 10.5.0, GraphPad Software, San Diego, CA, USA) [20] to create waterfall graphs and scatter plots, and Microsoft Excel to calculate z-scores. A 95% confidence interval (mean + 1.96 x SD) was determined, and the states that fell outside this range on the waterfall plots were considered statistically (p < .05) different [21]. States that fell outside an 86% confidence interval (+1.5 x SD) and the fold difference between the highest and lowest states were noted. Exploratory associations between hydromorphone use with state characteristics [17, 18] were determined with Pearson r. A R value of ±0.1 to ±0.3 accounts for a small, ±0.4 to ±0.6 ≅ medium, ±0.7 to ±0.9 ≅ large, and ±0.9 to ±1.0 ≅ very large portion of the variance [22].

## 3. Results

### 3.1. ARCOS 2010-2023

There was an increase in distribution from 2010 to 2013 (+30.6%), peaking at 1,839.45 kg. This was followed by a large decrease (−55.9%) by 2023, reaching a low of 810.94 kg. Overall, pharmacies accounted for the preponderance (79.8%) whereas hospitals (19.4%) and practitioners (0.8%) were responsible for a subset of distribution in 2023 (Figure 1).

**Figure 1:**
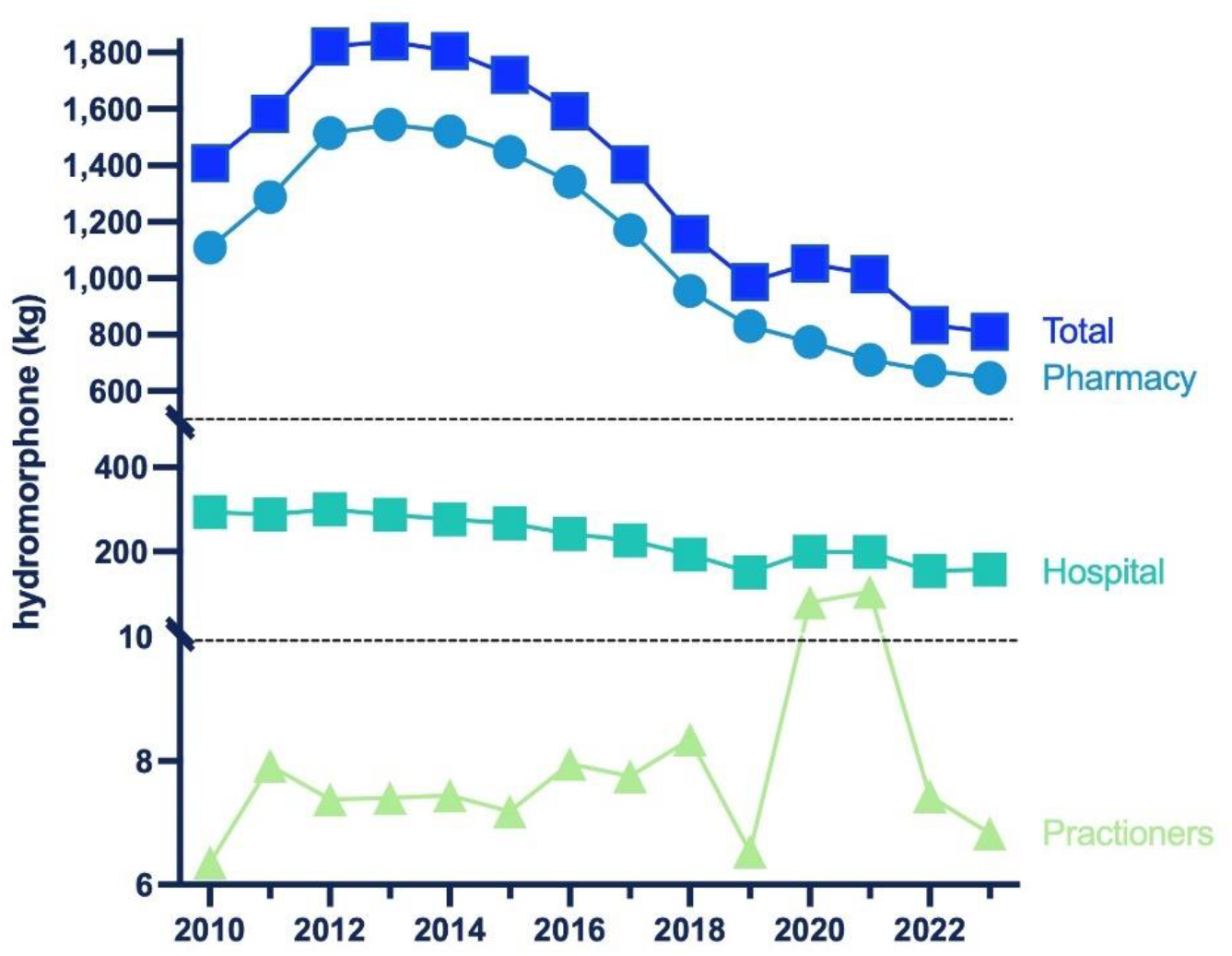
Hydromorphone in kilograms distributed by business activity from 2010 to 2023 reported by the US Drug Enforcement Administration’s Automated Reports and Consolidated Orders System (ARCOS).

The milligrams per person decreased in all but two states (NY and MS). Also, the Midwestern states (WI, ND) had a higher percentage decrease (Figure 2A). New York had the highest increase (+104.0%) while Wisconsin had the most decrease (−74.6%). There was a greater decrease (−84.2 to −71.8%) in the smaller cities whereas the least change (−47.0 to −2.0%) was in the bigger cities of New York. In Wisconsin, the metropolitan areas showed the most percent decrease (−84.2 to −71.8%), whereas the rural areas showed a lower percent change (−47.0 to −2.0%) (Supplemental Figure 1).

**Figure 2:**
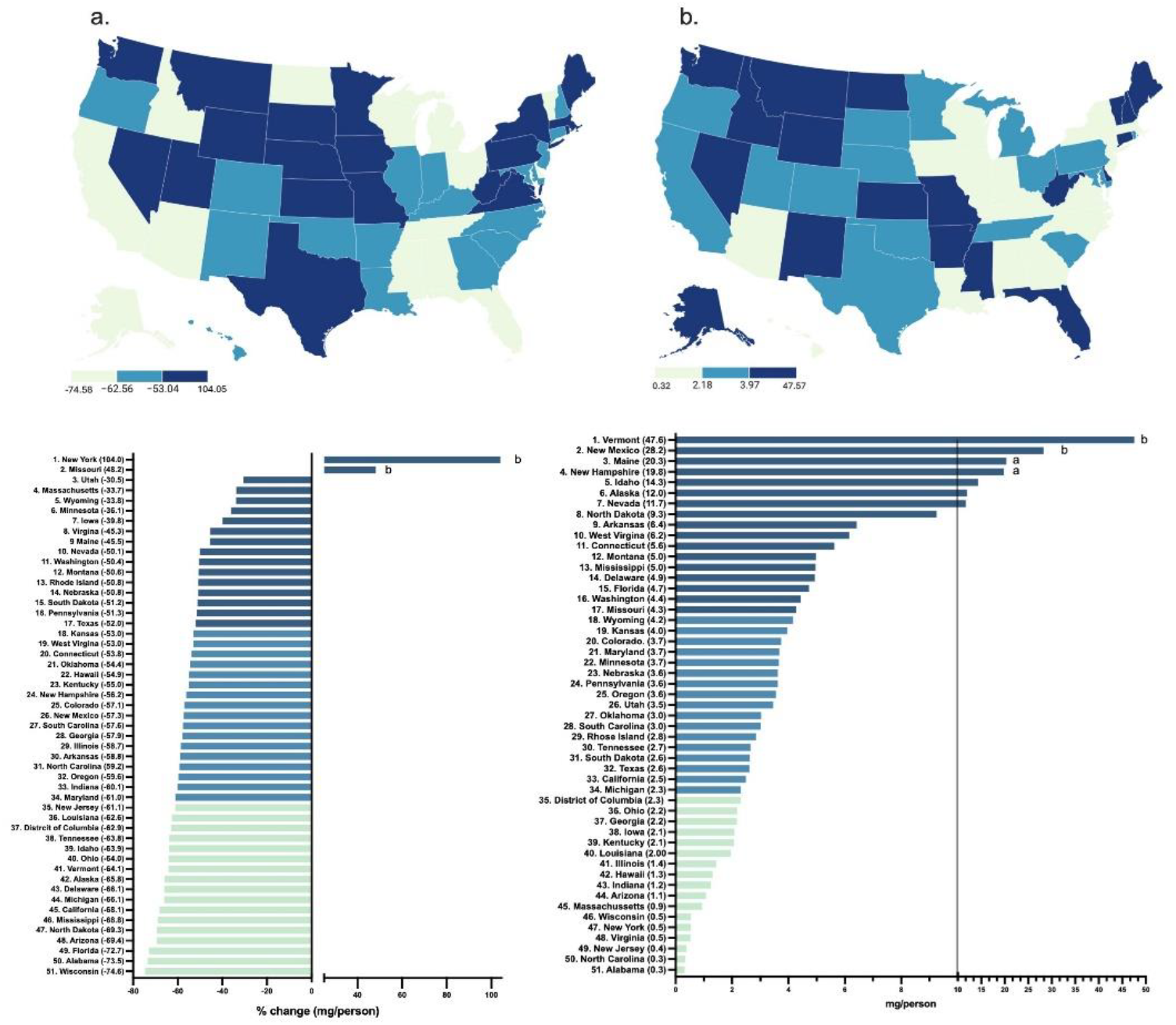
Drug Enforcement Administration’s Automated Reports and Consolidated Orders System (ARCOS) percent change in mg/person of hydromorphone from 2013 to 2023 (a) and mg/person in 2023 (b, outside of ^A^1.5 or ^B^1.96 SDs).

In 2023, Vermont had the highest milligrams per person (47.6, p < .05) whereas Alabama had the lowest (0.3). There was a 158.7-fold difference between them (Figure 2B).

### 3.2. Medicaid 2013-2023

Since 2013, there was an increase (+39.6%) in total prescriptions by 2015, peaking at 1,192,339. There was a large decrease (−48.9%) in prescriptions by 2023, reaching a low of 609,530 prescriptions. The brand name Exalgo had practically vanished by 2023 (accounting for 0.0% of the total), whereas Dilaudid accounted for almost one-fifth of prescriptions (19.3%). Overall, generic (>80%) stayed consistent and made up almost all prescriptions of hydromorphone (Figure 3).

**Figure 3:**
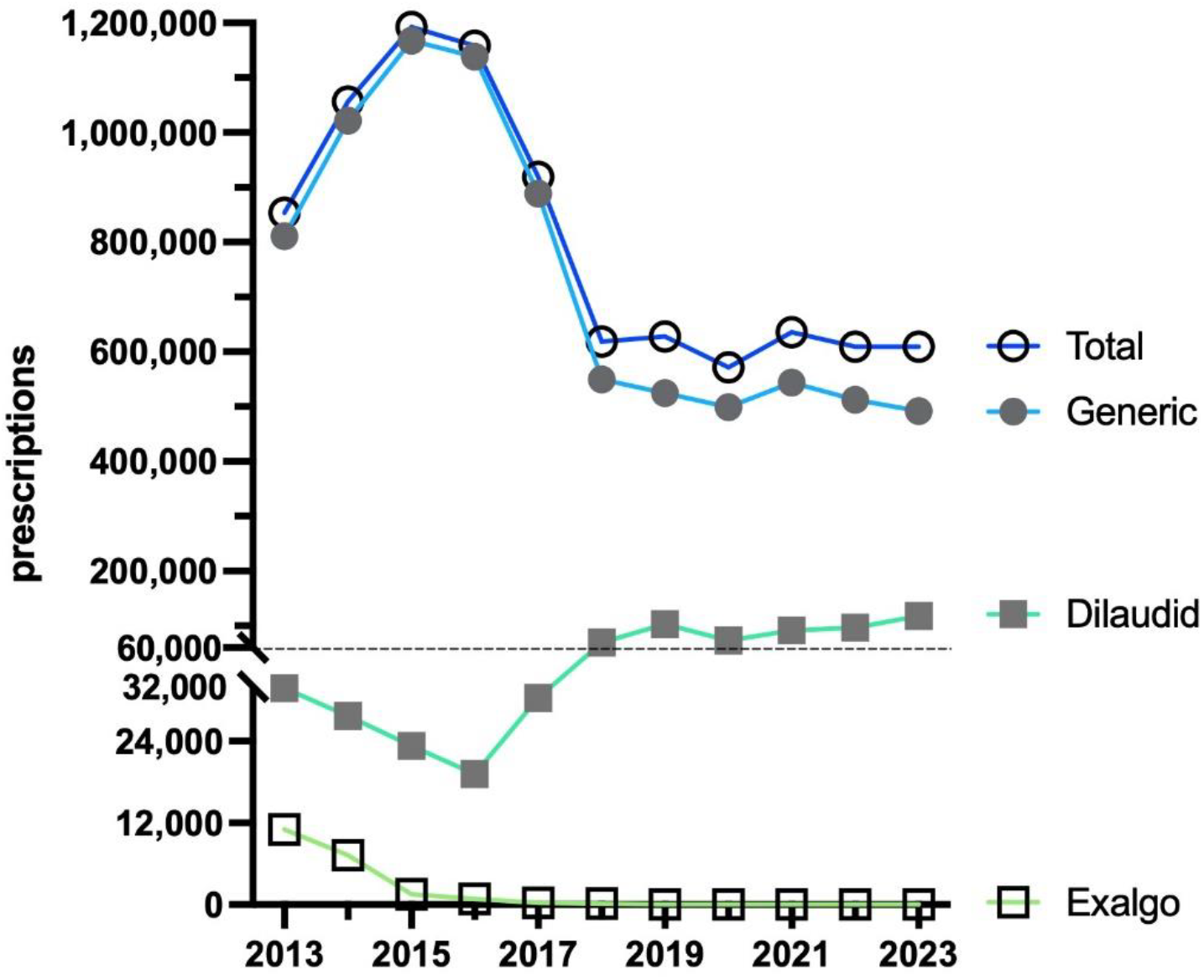
Prescriptions of hydromorphone to US Medicaid patients [9] by formulation from 2013 to 2023.

By 2023, the Medicaid prescriptions of hydromorphone decreased by a wide range in almost all states. Only a few states (MS, MA, ID, NE) showed a positive change. The Western states (WY, UT, CO) showed the most negative change, whereas the Midwestern states (IA, KS, SD) only had a small decrease (Figure 4a).

**Figure 4:**
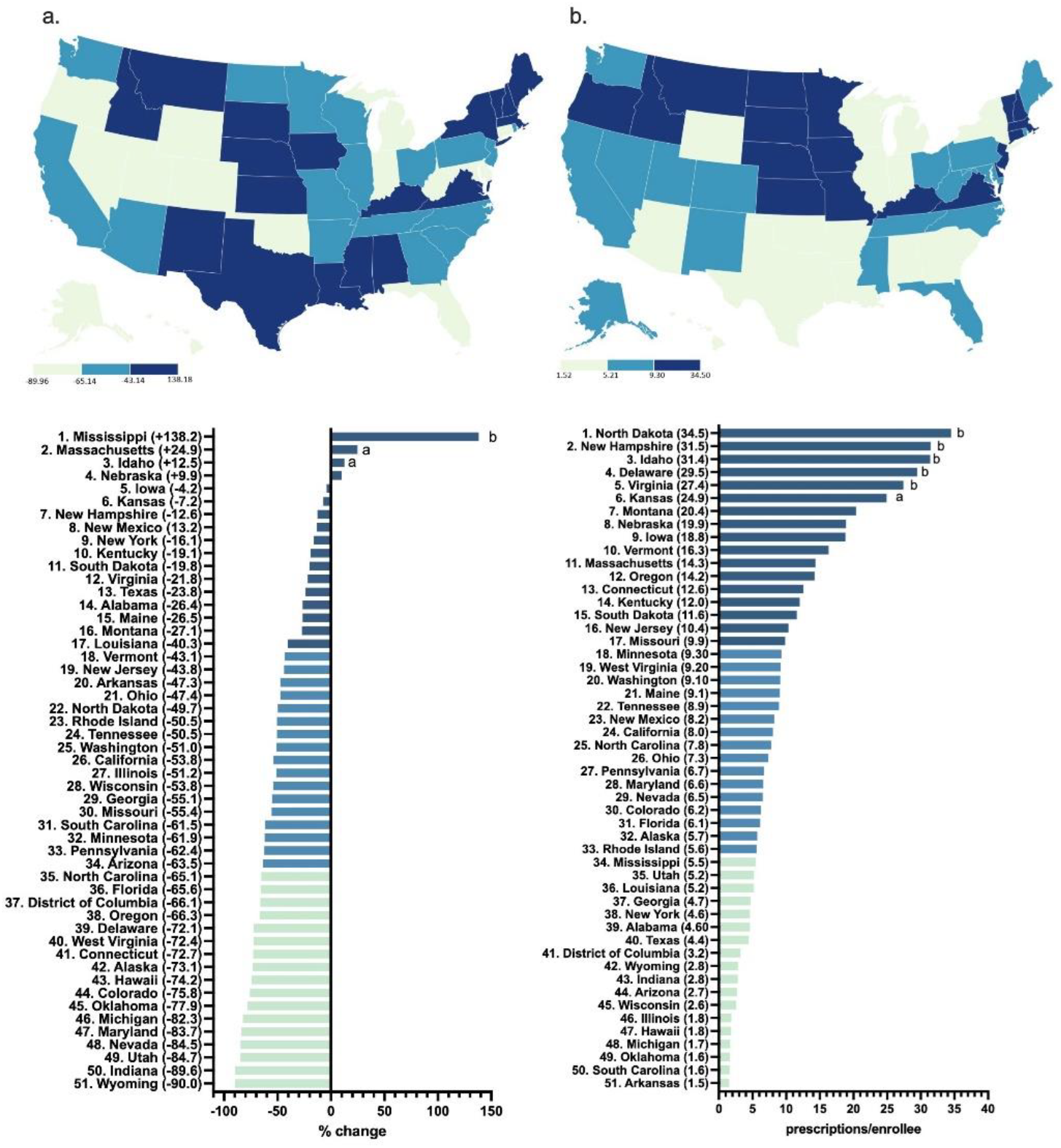
Percent change in US Medicaid prescriptions [9] per thousand enrollees from 2015 to 2023 (a) and prescriptions per thousand enrollees in 2023 (b, outside of ^A^1.5 or ^B^1.96 SDs).

In 2023, North Dakota had the highest prescriptions per enrollees hydromorphone (3.5) whereas Arkansas had the lowest (0.2), showing a 17.5-fold difference between them (Figure 4b).

### 3.3 Medicare 2013-2023

The data showcased an increase (+8.5%) in total Medicare claims by 2015, peaking at 1,308,316 claims. By 2023, there was a large decrease (−31.9%) in claims, reaching a low of 891,538. The brand names Dilaudid (0.2%) and Exalgo (0.0%) had almost vanished by 2023, whereas the generic (99.8%) stayed consistent with the total Medicare claims (Figure 5).

**Figure 5:**
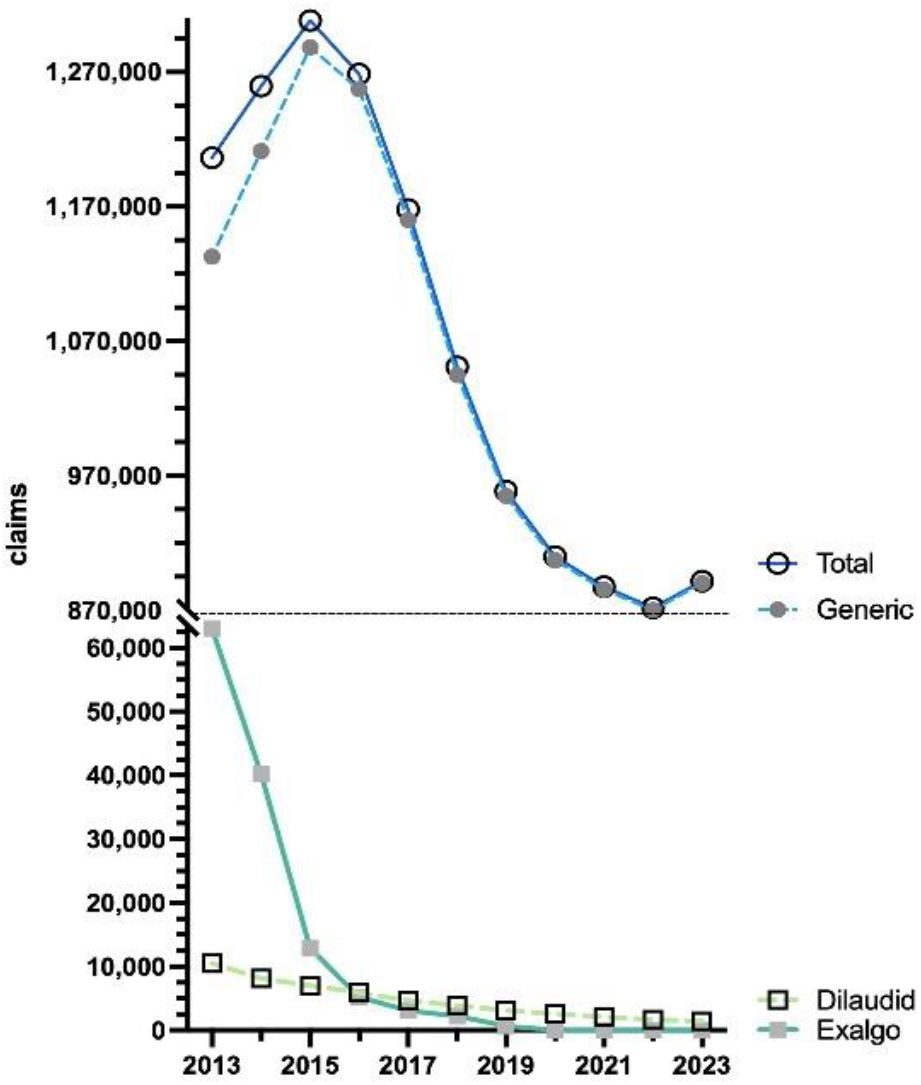
US Medicare Part D [10] claims of hydromorphone by formulation from 2013 to 2023.

By 2023, the claims increased by a wide range (+1.6 to 119.5%) in almost all states. Only a small subset of states (21) showed a negative change. The Northeastern states (MN, CT, RI) increased the most, whereas the Midwestern states (ND, WV, KS) decreased the most (Figure 6a).

**Figure 6:**
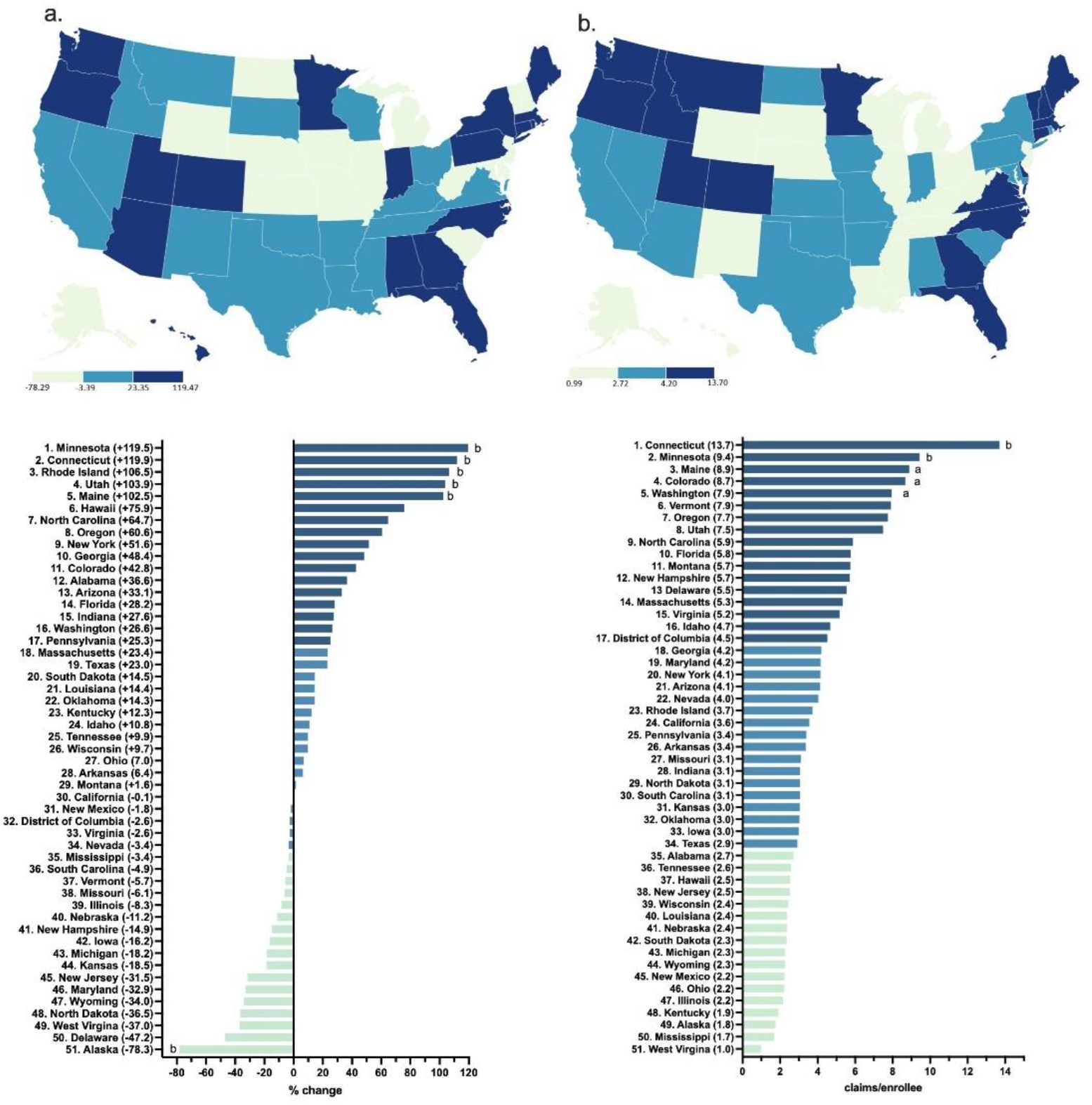
Percent change in US Medicare Part D claims [10] per enrollee from 2015 to 2023 (a) and claims per hundred enrollees of hydromorphone in 2023 (b, outside ^A^1.50 and ^B^1.96 SDs).

In 2023, Connecticut had the highest claims per enrollee hydromorphone (13.7) whereas West Virginia had the lowest (1.0), showing a 13.7-fold difference between them (Figure 6b).

### 3.4. Correlations

ARCOS in 2013 highly correlated with ARCOS in 2023 data (R^2^(49) = 0.970, p < 0.0001) (Figure 7a). Within Medicaid, the prescriptions in 2015 were highly associated with 2023 prescriptions (R^2^(49) = 0.422, p < 0.0001) (Figure 7b). In 2015, the Medicare claims were highly correlated to Medicaid prescriptions (R^2^(49) = 0.466, p < 0.0001) (Figure 7c). In 2023, the Medicaid prescriptions had a moderate negative correlation with the population demographics (% non-whites) (R^2^(49) = 0.128, p = 0.010) (Figure 7d) and the ARCOS distribution moderately correlated with the median ages (R^2^(49) = 0.107, p = 0.019) (Figure 7e). Also in 2023, the Medicare claims had a small, but significant, association with providers per thousand population (R^2^(49) = 0.085, p = 0.038) (Figure 7f). All other correlations between the ARCOS distribution, Medicaid prescriptions, Medicare claims, and state demographics are presented in Table 2.

**Table 2.**
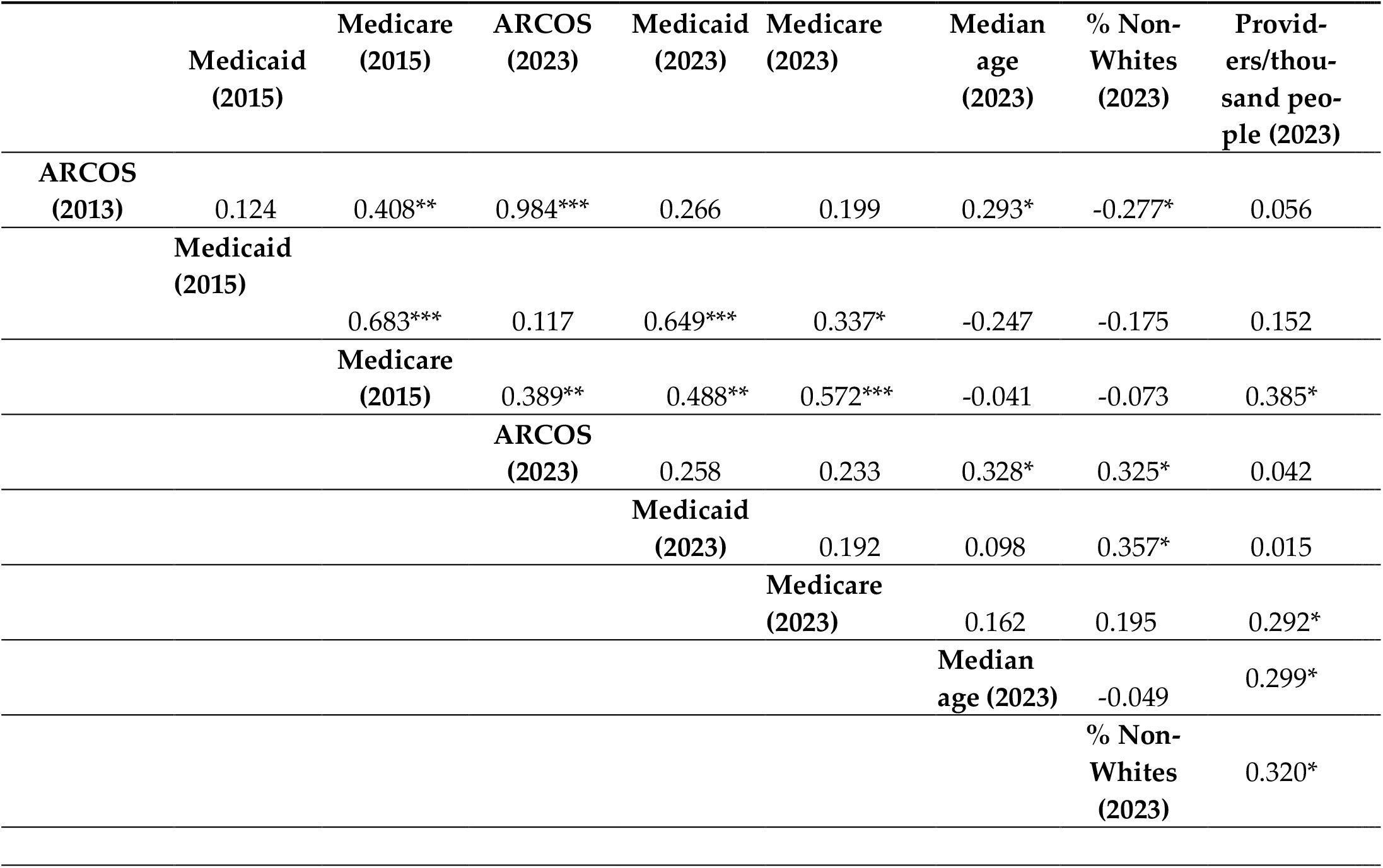
ARCOS, ARCOS, Medicaid, and Medicare correlations with state level demographics including median age [14], percent non-White [18], providers per thousand population [17]. (* p < .05, **p < .005, and *** p< .0001).

|  | Medicaid<br>(2015) | Medicare<br>(2015) | ARCOS<br>(2023) | Medicaid<br>(2023) | Medicare<br>(2023) | Median<br>age<br>(2023) | % Non-Whites<br>(2023) | Provid-<br>ers/thou-<br>sand peo-<br>ple (2023) |
| --- | --- | --- | --- | --- | --- | --- | --- | --- |
| **ARCOS**<br>**(2013)** | 0.124 | 0.408** | 0.984*** | 0.266 | 0.199 | 0.293* | -0.277* | 0.056 |
| **Medicaid**<br>**(2015)** |  | 0.683*** | 0.117 | 0.649*** | 0.337* | -0.247 | -0.175 | 0.152 |
|  |  | **Medicare**<br>**(2015)** | 0.389** | 0.488** | 0.572*** | -0.041 | -0.073 | 0.385* |
|  |  |  | **ARCOS**<br>**(2023)** | 0.258 | 0.233 | 0.328* | 0.325* | 0.042 |
|  |  |  | **Medicaid**<br>**(2023)** |  | 0.192 | 0.098 | 0.357* | 0.015 |
|  |  |  |  | **Medicare**<br>**(2023)** |  | 0.162 | 0.195 | 0.292* |
|  |  |  |  |  | **Median**<br>**age (2023)** |  | -0.049 | 0.299* |
|  |  |  |  |  |  |  | **% Non-**<br>**Whites**<br>**(2023)** | 0.320* |

**Figure 7:**
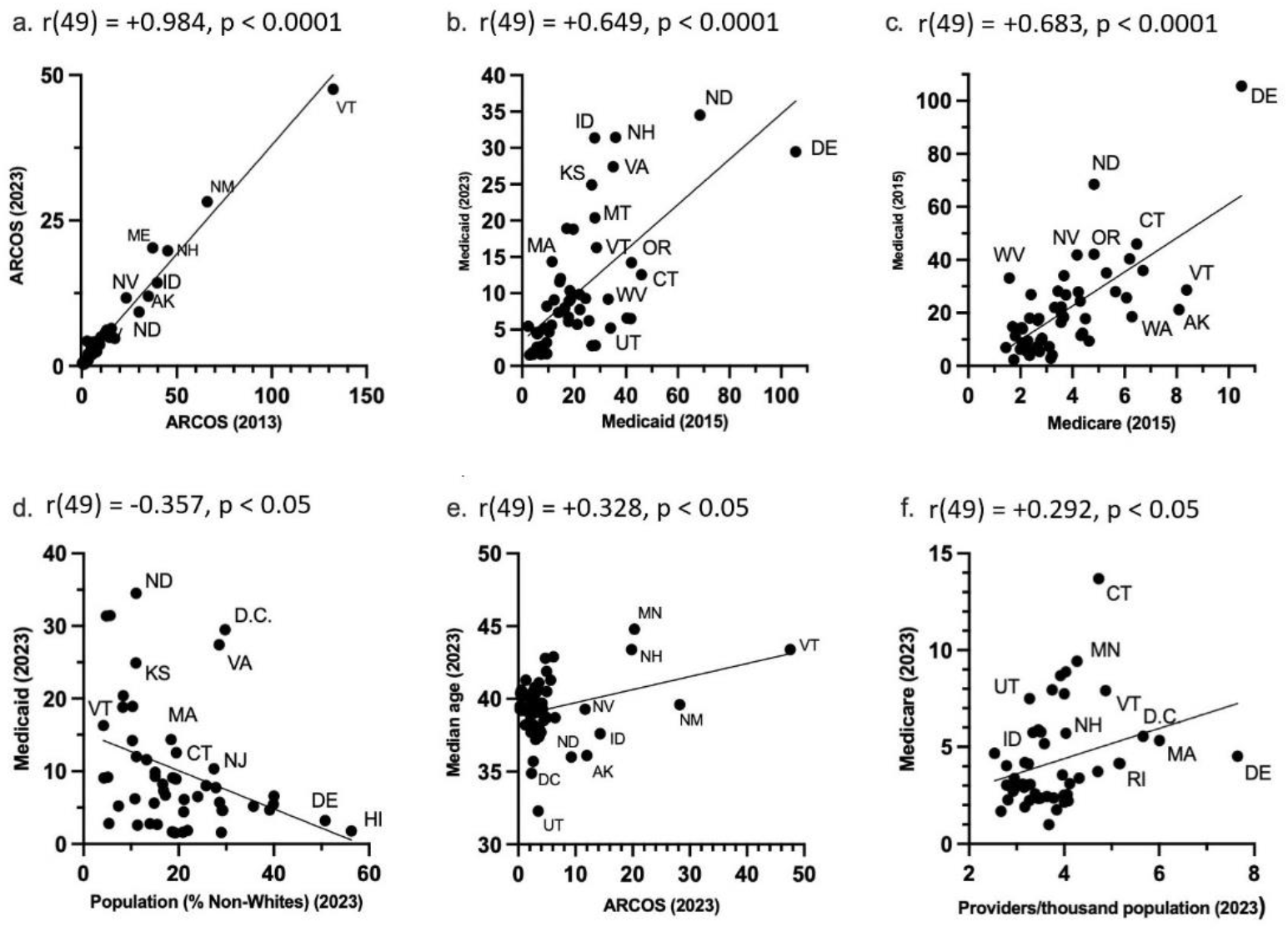
State-level ARCOS, Medicaid, and Medicare scatter plots. (a). ARCOS 2013 mg/person correlation with ARCOS 2023 mg/person. (b). Medicaid 2015 prescriptions/thousand enrollees correlated with Medicaid 2023 prescriptions/thousand enrollees. (c). Medicare 2015 claims/hundred enrollees correlated with Medicaid 2015 prescriptions/ thousand enrollees. (d). 2023 Population percentage Non-Whites correlated with Medicaid 2023 prescriptions/ thousand enrollees. (e). ARCOS 2023 mg/person correlated with 2023 population median age. (f). 2023 Providers/ thousand population correlated with Medicare 2023 claims/hundred enrollees.

## 4. Discussion

This novel study identified regionally dependent disparities in hydromorphone use in the US between 2010 to 2023. According to ARCOS data, hydromorphone distribution increased by +30.6% through 2013, followed by a decrease of −55.9% by 2023 (Figure 1). A similar pattern was observed in both Medicaid and Medicare programs. Medicaid hydro-morphone prescriptions increased by +39.6% by 2015, then decreased by −48.9% by 2023 (Figure 3). Similarly, Medicare Part D hydromorphone claims increased by +8.5% by 2015, then decreased by −31.9% by 2023 (Figure 5). Given that hydromorphone is typically prescribed only when initial treatments have proven ineffective, primarily due to the drug’s elevated potency, potential for abuse, and risk of overdose [23], there is a critical need to better understand this usage pattern.

Since 2000, there has been a more than 1,000% increase in opioid overdose deaths [24]. In 2021, an estimated 2.5 million people aged 18 or older had opioid use disorder, yet only about one in five received treatment [25]. By December 2023, there were 20% fewer deaths than there were in December 2022 [26]. Overall, while opioid-related deaths may be declining, the ongoing opioid crisis is far from over.. In 2023, nearly 110,037 people in the United States died from drug overdoses, with approximately 83,140 deaths involving an opioid [25]. A recent study among 29 states and the District of Columbia showed that the percentage of overdose deaths involving counterfeit pills more than doubled from July 2019 to December 2021. Since then, many policy interventions have targeted the opioid overdose epidemic [27]. “Overdose Data to Action supports jurisdictions in implementing prevention activities and in collecting accurate, comprehensive, and timely data on nonfatal and fatal overdoses and in using those data to enhance programmatic and surveillance efforts” [28]. The Opioid Rapid Response Program is a federal effort to reduce opioid overdose deaths. The program helps address care continuity and risk reduction for patients by alerting state health agencies about federal law enforcement events that might disrupt patients’ access to care and supporting state and local capacity building to prepare for and respond to disruptions in care [29]. In addition to these policies, states have adopted Prescription Drug Monitoring Programs (PDMPs), which are electronic databases that track controlled substance prescriptions [30]. Information from PDMPs can help clinicians identify patients who may be at risk for overdose and provide potentially lifesaving information and interventions [31]. Lastly, the Centers for Medicare and Medicaid Services implemented new coverage policies that now ensure that starting January 1, 2020, Medicare covers methadone for MAT and related services furnished by opioid treatment programs (OTPs) [32]. The decline of hydromorphone use suggests that these policies may have been effective in reducing access to the opioid as part of efforts to control the opioid crisis in the US [33].

Additionally, it is important to note that the hydromorphone aggregate production quota decreased by −71.5% from 2015 to 2023. The DEA’s quota is adjusted “in order to provide for the estimated medical, scientific, research, and industrial needs of the U.S., lawful export requirements, and the establishment and maintenance of reserve stocks” [34]. Our data suggests that the DEA’s aggregate production quota decrease was probably a factor in the slow decline of hydromorphone distribution from 2010 to 2023 in the US as well.

Importantly, hydromorphone was distributed primarily to pharmacies (79.8%), with hospitals (19.4%) and practitioners (0.8) being a small subset of the total distribution in 2023 (Figure 1). Nearly 7,000 pharmacies have closed since 2019. Many community pharmacies are struggling to stay open due to an overburdened workforce, shrinking reimbursement rates for prescription drugs and limited opportunities to bill insurers for services beyond dispensing medications [35]. The closure of pharmacies may account for the decline in the hydromorphone distributions shown in the ARCOS data.

By 2023, New York state had an increase in hydromorphone distribution of +104.0%, with the elevation occurring largely in the state’s major metropolitan areas. However, Wisconsin had the highest decrease of −74.6%, with the reduction also concentrated primarily in the metropolitan areas of the state (Supplemental Figure 1). These disparities between states showcase that there might be something more than just the pharmacy closures driving the decline of hydromorphone. The population of New York is increasing rapidly every day, reaching a high of 15,611,308 adults in 2023 [36]. Notably, since the data is corrected by population with milligrams of hydromorphone per adult in New York, the increase in the population drives the elevation in New York’s distribution as well as the regional increase in the larger, more populated, cities in the Empire State. Additionally, there were pronounced regional disparities in hydromorphone use identified in ARCOS (158.7 fold), Medicaid (17.5 fold), and Medicare Part D (13.7 fold) (Figures 2b,4b,6b). Medicaid prescriptions per thousand enrollees of hydromorphone had decreased in all but four states (MS, MA, ID, NE) from 2015 to 2023. Regionally, the Western states like Wyoming, Utah, and Colorado had the most negative change, whereas the Midwestern states like Iowa, Kansas, and South Dakota only had a small negative change (Figure 4a). Regionally, the Northeastern states like Minnesota, Connecticut, and Rhode Island increased the most, whereas Midwestern states like North Dakota, West Virgina, and Kansas decreased the most (Figure 6a).

Even though nationally the overall Medicare Part D claims for hydromorphone decreased, the per capita (claims/thousand enrollee) for each state actually increased by a wide range in most states from 2015 to 2023. According to the CMC’s Medicare Part D prescribing guide, all Part D plans have a DMP that limits access to opioids that are frequently abused. This includes the seven-day supply limit alert that “limits initial opioid fills for Part D patients who haven’t filled an opioid prescription recently, such as within the past 60 days, to a supply of 7 days or less. [37] This alert shouldn’t affect patients who already take opioids.” This suggests that the access to opioids is tightened for new patients, but the original patients who already receive the prescriptions for opioids are not affected. For example, some states might have prescribed hydromorphone to fewer patients, but those who did might have received the drug in more concentrated and more frequent prescriptions. This would explain why overall the hydromorphone Medicare claims in the US decreased from 2015 to 2023, but the per capita claims increased in most states.

These three databases were correlated with each other and state population demographics. The 2013 ARCOS very highly correlated (r = 0.984) with the 2023 ARCOS distribution (Figure 7a). This indicated a long-term consistency in the distribution of hydromorphone. Similarly, there was a moderate correlation (r = 0.649) between the 2015 Medicaid and the 2023 Medicaid prescriptions (Figure 7b). This indicated persistent state-level differences in the Medicaid prescriptions over time. There was also a high correlation (r = 0.683) between the Medicaid 2015 data and Medicare 2023 data (Figure 7c). This indicated some level of shared prescribing environment or policies between the two public healthcare programs as well.

Additionally, there was a negative moderate association (r = −0.357) between the Medicaid prescriptions with the Non-White population per state in 2023 (Figure 7d). This indicated the racial disparity in access to the opioid, where states with higher Non-White populations had lower Medicaid prescriptions. The CDC’s overdose prevention website states that the percentage receiving treatment in the US was lowest for Black, Hispanic, and American Indian or Alaska Native racial and ethnic groups [38]. This would also explain the racial disparity seen in the Medicaid 2023 prescriptions. There was also a low correlation (r = 0.328) between the ARCOS distribution and the average median ages per state in 2023 (Figure 7e). This suggested the age-related needs of hydromorphone in states where the median age was higher. The CDC stated in a recent data brief that in 2023, 24.3% of adults had chronic pain, and 8.5% of adults had chronic pain that also increases with age [39]. This would make sense as to why hydromorphone distribution is higher in states with higher median ages as well. Lastly, there was a low correlation (r = 0.292) between the Medicare 2023 data and the providers per thousand population per state in 2023. This suggested an access related disparity where states with lower number of providers had lower Medicare claims. The Association of American Medical Colleges predicted that the United States will face a physician shortage of up to 86,000 physicians by 2036 [40]. The Kaiser Family Foundation also stated that nearly three million Americans live in rural areas that lack both health care and internet, which in turn create access barriers for the patients [41]. This also provides a possible explanation as to why there are fewer hydromorphone claims in the states with lower number of providers.

These persistent and significant state-level disparities suggest that regional prescribing cultures and healthcare patterns continue to influence the geographic variation in opioid prescribing. These findings stress that for opioid policies there is no one size fit all, and that many states and regions of the US are influenced by other outcomes like access, population, and demographics as well. The clinical utility of hydromorphone is inherently limited by its pharmacological profile. While its high lipophilicity and rapid onset make it effective for severe pain management, these same properties contribute to its high abuse potential and overdose risk. The drug’s primary FDA indication for use, “for the management of pain severe enough to require an opioid analgesic and for which alternate treatments are inadequate only when other treatments have failed” reflects appropriate clinical caution. However, the historical prescribing patterns and state-level variation revealed in the study suggest this restriction may not have been consistently applied. The availability of multiple formulations (oral, parenteral) and the drug’s ability to cross the blood-brain barrier more readily than morphine underscore its utility for pain management while at the same time the need for careful patient selection and monitoring [42, 43, 44].

A limitation that comes with the ARCOS database is that it does not provide any information on how many people are receiving hydromorphone or who is getting it and simply provides the weight of the drug distributed to each state. However, Medicaid and Medicare Part D programs fills that gap by providing information on the number of prescriptions and claims for hydromorphone. This extends previous reports that looked at only distribution [7]. The correlations between the three databases suggest that the lack of specificity in ARCOS was in fact captured by Medicaid and Medicare. Therefore, future studies can focus on looking at individual patient data to understand the disparities at a patient level in addition to the ecological disparities found in this study. For the future, it is important that interventions and policies are regionally tailored to account for the demographic and geographical differences that impact not only the access to medications, but also healthcare in general.

Several state-level disparities were shown through this study, with a decline in hydromorphone use being the primary finding. Disparities that include varying levels of data availability, opioid use, and mortality rates were prevalent across the different states. This complexity is compounded by a rise in chronic pain conditions, underemphasized education for health care providers in pain management, and a limited number of specialists in pain management and addiction medicine. To combat these disparities, the American Hospital Association has implemented Opioid stewardship programs that promote appropriate use of opioid medications, improve patient outcomes and reduce misuse of opioids [45]. These programs will continue to focus on prescription opioids in the future to reduce state level disparities and declines that have been observed through this study.

## 5. Conclusions

This multi-database analysis demonstrates that hydromorphone utilization in the United States has undergone a significant transformation over the past decade, with a marked decline following peak usage around 2015. The convergent findings across ARCOS, Medicaid, and Medicare data sources provide robust evidence that policy interventions and clinical guideline changes may have successfully reduced access to this high-potency opioid. However, persistent regional disparities highlight the need for continued surveillance and targeted interventions to ensure appropriate pain management while minimizing opioid-related harms.

## Supporting information

Supplemental Figure 1:ARCOS Percent change mg/person of hydromorphone from 2013 to 2023 by zip-code in New York and Wisconsin.

## Data Availability

All data produced in the present study are contained in the manuscript.

## Supplementary Materials

The following supporting information can be downloaded at: https://www.mdpi.com/article/doi/s1, Figure S1: Drug Enforcement Administration’s Automated Reports and Consolidated Orders System (ARCOS). Percent change mg/person of hydromorphone from 2013 to 2023 by zip-code in New York and Wisconsin.

## Author Contributions

Conceptualization, BJP. And KSP.; methodology, BJP.; software, KSP.; validation, BJP, KSP and KLM; formal analysis, KSP.; investigation, KSP.; resources, KSP.; data curation, KSP.; writing—original draft preparation, KSP.; writing—review and editing, BJP, KLM.; visualization, KSP.; supervision, BJP, KLM.; project administration, BJP.; funding acquisition, BJP. All authors have read and agreed to the published version of the manuscript.

## Institutional Review Board Statement

**The study was conducted in accordance with the Declaration of Helsinki, and approved by the Institutional Review Board of Geisinger**.

## Informed Consent Statement

Not applicable.

## Data Availability Statement

Raw data is available [8,9,10}. Extracted data (add link currently being generated).

## Acknowledgments

Thank you to the Summer Undergraduate Research Program for their support.

## Abbreviations

The following abbreviations are used in this manuscript:

ARCOS: Drug Enforcement Administration’s Automated Reports and Consolidated Orders System
CDC: Centers for Disease Control and Prevention
CMS: Centers of Medicare and Medicaid Services
US: United States

## Disclaimer/Publisher’s Note

The statements, opinions and data contained in all publications are solely those of the individual author(s) and contributor(s) and not of MDPI and/or the editor(s). MDPI and/or the editor(s) disclaim responsibility for any injury to people or property resulting from any ideas, methods, instructions or products referred to in the content.

## References

1. .Dowell D, Ragan KR, Jones CM, Baldwin GT, Chou R. CDC Clinical Practice Guideline for Prescribing Opioids for Pain - United States, 2022. MMWR Recomm Rep. 2022 Nov 04;71(3):1–95

2. Liu L, Xu M, Wang J, Hu Y, Huang Z. Research Progress of Hydromorphone in Clinical Application. Physiological Research [Internet]. 2025 Mar 10;(1/2025):41–8. Available from: https://www.biomed.cas.cz/physiolres/pdf/74/74_41.pdf

3. Murray A, Hagen NA. Hydromorphone. Journal of Pain and Symptom Management [Internet]. 2005 May 1;29(5):57–66. Available from: https://www.jpsmjournal.com/article/S0885-3924(05)00033-3/fulltext

4. Dowell D, Ragan K, Jones C, Baldwin G, Chou R. CDC Clinical Practice Guideline for Prescribing Opioids for Pain. MMWR Recommendations and Reports [Internet]. 2022 Nov 4;71(3):1–95. Available from: https://www.cdc.gov/mmwr/vol-umes/71/rr/rr7103a1.htm

5. U.S. Drug Enforcement Administration, Diversion Control Division. Hydromorphone. Washington (DC): DEA; 2024 Feb. Available from: https://deadiversion.usdoj.gov/drug_chem_info/hydromorphone.pdf

6. Mutter R, Black J, Iwanicki J. Changes in the street prices of prescription opioids during the COVID-19 pandemic. Psychiatr Serv. 2023;74(1):63–5. doi:10.1176/appi.ps.202100689.

7. Madera JD, Ruffino AE, Feliz A, McCall KL, Davis CS, Piper BJ. Declining but Pronounced State-Level Disparities in Prescription Opioid Distribution in the United States. Pharmacy. 2024 Jan 16;12(1):14–4.

8. Drug Enforcement Administration. ARCOS Retail Drug Summary Reports [Internet]. Washington (DC): U.S. Departmentof Justice; [cited 2025 Jul 3]. Available from: https://www.deadiversion.usdoj.gov/arcos/retail_drug_summary/arcos-drug-summary-reports.html

9. Medicaid.gov. State Drug Utilization Data [Internet]. Baltimore (MD): Centers for Medicare & Medicaid Services; [cited 2025 Jul 3]. Available from: https://www.medicaid.gov/medicaid/prescription-drugs/state-drug-utilization-data

10. Centers for Medicare & Medicaid Services. Medicare Part D Prescribers - by Geography and Drug [Internet]. Baltimore (MD): CMS; [cited 2025 Jul 3]. Available from: https://data.cms.gov/provider-summary-by-type-of-service/medicare-part-d-prescribers/medicare-part-d-prescribers-by-geography-and-drug

11. MedlinePlus. Hydromorphone: MedlinePlus Drug Information [Internet]. Bethesda (MD): U.S. National Library of Medicine; [updated 2024 May 15; cited 2025 Jul 11]. Available from: https://medlineplus.gov/druginfo/meds/a682013.html

12. Khadivi A, Hughes B. Hydromorphone. In: StatPearls [Internet]. Treasure Island (FL): StatPearls Publishing; 2024 Jan– [cited 2025 Jul 11]. Available from: https://www.ncbi.nlm.nih.gov/books/NBK470393/

13. U.S. Drug Enforcement Administration, Diversion Control Division. Hydromorphone [Internet]. Springfield (VA): DEA; 2024 Feb [cited 2025 Jul 11]. Available from: https://deadiversion.usdoj.gov/drug_chem_info/hydromorphone.pdf

14. U.S. Census Bureau. State population totals and components of change: 2020–2024 [Internet]. Washington (DC): U.S. Census Bureau; 2025 May 28 [cited 2025 Jul 3]. Available from: https://www.census.gov/data/tables/time-se-ries/demo/popest/2020s-state-total.html

15. Medicaid.gov. State Drug Utilization Data Datasets [Internet]. Baltimore (MD): Centers for Medicare & Medicaid Services; [cited 2025 Jul 3]. Available from: https://data.medicaid.gov/dataset/6c114b2c-cb83-559b-832f-4d8b06d6c1b9

16. Centers for Medicare & Medicaid Services. Monthly Enrollment by State [Internet]. Baltimore (MD): CMS; [cited 2025 Jul 3]. Available from: https://www.cms.gov/data-research/statistics-trends-and-reports/medicare-advantagepart-d-contract-and-enrollment-data/monthly-enrollment-state

17. Association of American Medical Colleges. U.S. Physician Workforce Data Dashboard [Internet]. Washington (DC): AAMC; [cited 2025 Jul 3]. Available from: https://www.aamc.org/data-reports/report/us-physician-workforce-data-dash-board

18. U.S. Census Bureau. State population estimates: 2020s [Internet]. Washington (DC): U.S. Census Bureau; [cited 2025 Jul 3]. Available from: https://www.census.gov/data/datasets/time-series/demo/popest/2020s-state-detail.html

19. Datawrapper. Datawrapper: Create charts, maps, and tables [Internet]. Berlin: Datawrapper GmbH; [cited 2025 Jul 3]. Available from: https://www.datawrapper.de/

20. Berkman, S.J., Roscoe, E.M. and Bourret, J.C. (2019), Comparing self-directed methods for training staff to create graphsusing Graphpad Prism. Jnl of Applied Behav Analysis, 52: 188–204. 10.1002/jaba.522

21. Eidbo, S. A., Kropp Lopez, A. K., Hagedorn, J. D., Mathew, V., Kaufman, D. E., Nichols, S. D., McCall, K. L., & Piper, B. J. (2022). Declines and regional variation in opioid distribution by U.S. hospitals. Pain, 163(6), 1186–1192. 10.1097/j.pain.0000000000002473

22. Akoglu H. User’s Guide to Correlation Coefficients. Turkish Journal of Emergency Medicine [Internet]. 2018;18(3):91–3. Available from: https://pmc.ncbi.nlm.nih.gov/articles/PMC6107969/

23. Dowell D, Ragan K, Jones C, Baldwin G, Chou R. CDC Clinical Practice Guideline for Prescribing Opioids for Pain. MMWR Recommendations and Reports [Internet]. 2022 Nov 4;71(3):1–95. Available from: https://www.cdc.gov/mmwr/vol-umes/71/rr/rr7103a1.htm

24. NIHCM Foundation. Visualizing the Impact of the Opioid Overdose Crisis [Internet]. Washington (DC): National Institute for Health Care Management; 2022 Jul 21 [cited 2025 Jul 21]. Available from: https://nihcm.org/publications/visualizing-the-impact-of-the-opi-oid-overdose-crisis

25. GovFacts. The Opioid Crisis: Prevention, Treatment, and Recovery [Internet]. GovFacts; [updated 2025 Jul; cited 2025 Jul 21]. Available from: https://govfacts.org/federal/cdc/the-opioid-crisis-prevention-treatment-and-recovery/

26. Saunders H, Panchal N, Zitter S. Opioid Deaths Fell in Mid-2023, But Progress Is Uneven and Future Trends Are Uncertain [Internet]. San Francisco (CA): KFF; 2024 Sep 23 [cited 2025 Jul 21]. Available from: https://www.kff.org/mental-health/issue-brief/opioid-deaths-fell-in-mid-2023-but-progress-is-uneven-and-future-trends-are-uncertain/

27. Centers for Disease Control and Prevention (CDC). Overdose Prevention: Prevention Strategies [Internet]. Atlanta (GA): CDC; [updated 2025 Jun 9; cited 2025 Jul 21]. Available from: https://www.cdc.gov/overdose-prevention/prevention/index.html

28. Centers for Disease Control and Prevention (CDC). Overdose Data to Action (OD2A): About the Program [Internet]. Atlanta (GA): CDC; [cited 2025 Jul 21]. Available from: https://www.cdc.gov/overdose-prevention/php/od2a/about.html

29. Centers for Disease Control and Prevention (CDC). Opioid Rapid Response Program (ORRP) [Internet]. Atlanta (GA): CDC; [cited 2025 Jul 21]. Available from: https://www.cdc.gov/overdose-prevention/orrp/index.html

30. PDMP Training and Technical Assistance Center. Prescription Drug Monitoring Program (PDMP) Overview [Internet]. Brandeis University; [cited 2025 Jul 21]. Available from: https://www.pdmpassist.org/

31. Centers for Disease Control and Prevention (CDC). Prescription Drug Monitoring Programs (PDMPs) [Internet]. Atlanta (GA): CDC; 2024 May 7 [cited 2025 Jul 21]. Available from: https://www.cdc.gov/overdose-prevention/php/interventions/prescription-drug-moni-toring-programs.html

32. Centers for Medicare & Medicaid Services (CMS). Opioid Crisis in America [Internet]. Baltimore (MD): CMS; [cited 2025 Jul 21]. Available from: https://www.cms.gov/priorities/key-initiatives/opioids

33. Low CY, McCall KL, Piper BJ. Declines in Tapentadol Use in the US but Pronounced Regional Variation. Pharmacy (Basel, Switzerland) [Internet]. 2025 Summer;13(3):67. Available from: https://pubmed.ncbi.nlm.nih.gov/40407505/

34. Drug Enforcement Administration (DEA). Quotas for Controlled Substances [Internet]. Washington (DC): DEA Diversion Control Division; 2024 Dec 17 [cited 2025 Jul 21]. Available from: https://www.deadiversion.usdoj.gov/quotas/quotas.html

35. Berenbrok LA, Murphy M, Herbert S. Community Pharmacies Are Closing. Here’s What to Do If Your Neighborhood Location Does Too [Internet]. Arlington (VA): PBS NewsHour; 2025 Feb 22 [cited 2025 Jul 21]. Available from: https://www.pbs.org/news-hour/health/community-pharmacies-are-closing-heres-what-to-do-if-your-neighborhood-location-does-too

36. United States Census Bureau. State Population Totals and Components of Change: 2020–2024 [Internet]. Washington (DC): U.S. Department of Commerce; 2025 May 28 [cited 2025 Jul 21]. Available from: https://www.census.gov/data/tables/time-se-ries/demo/popest/2020s-state-total.html

37. Centers for Medicare & Medicaid Services (CMS). A Prescriber’s Guide to Medicare Prescription Drug (Part D) Opioid Policies [Internet]. Baltimore (MD): CMS; 2024 Aug [cited 2025 Jul 21]. Available from: https://www.cms.gov/files/document/mln2886155-pre-scribers-guide-medicare-prescription-drug-part-d-opioid-policies.pdf

38. Centers for Disease Control and Prevention (CDC). Achieving Health Equity Around Overdoses [Internet]. Atlanta (GA): CDC; 2024 May 8 [cited 2025 Jul 21]. Available from: https://www.cdc.gov/overdose-prevention/health-equity/achieving-health-equity-around-overdoses.html

39. Lucas JW, Sohi I. Chronic Pain and High-impact Chronic Pain in U.S. Adults, 2023 [Internet]. Hyattsville (MD): National Center for Health Statistics; 2024 Nov [cited 2025 Jul 21]. Available from: https://www.cdc.gov/nchs/products/databriefs/db518.htm

40. Association of American Medical Colleges (AAMC). New AAMC Report Shows Continuing Projected Physician Shortage [Internet]. Washington (DC): AAMC; 2024 Mar 21 [cited 2025 Jul 21]. Available from: https://www.aamc.org/news/press-releases/new-aamc-report-shows-continuing-projected-physician-shortage

41. Jane S. Millions in US Live in Places Where Doctors Don’t Practice and Telehealth Doesn’t Reach - KFF Health News [Internet]. KFF Health News. 2025. Available from: https://kffhealthnews.org/news/article/dead-zone-sickest-counties-slow-internet-broadband-de-sert-health-care-provider-shortage/

42. Murray A, Hagen NA. Hydromorphone. J Pain Symptom Manage. 2005 May;29(5 Suppl):S57–66.

43. Sarhill N, Walsh D, Nelson KA. Hydromorphone: pharmacology and clinical applications in cancer patients. Support Care Cancer. 2001 Mar;9(2):84–96.

44. U.S. Food and Drug Administration. Hydromorphone hydrochloride, USP [package insert]. Silver Spring, MD: FDA; 2023. Available from: https://www.accessdata.fda.gov/drugsatfda_docs/label/2023/019034s046lbl.pdf

45. American Hospital Association (AHA). *Stem the Tide: Opioid Stewardship Measurement Implementation Guide* [Internet]. Chicago (IL): AHA; 2020 May [cited 2025 Jul 21]. Available from: https://www.aha.org/system/files/media/file/2020/07/HIIN-opioid-guide-0520.pdf

