## Supplementary figures and images for "From Peak to Plunge: A Multi-Database Analysis of State Level Disparities in Hydromorphone use in the U.S. (2010-2023)"

### Supplemental Figure 1:ARCOS Percent change mg/person of hydromorphone from 2013 to 2023 by zip-code in New York and Wisconsin.

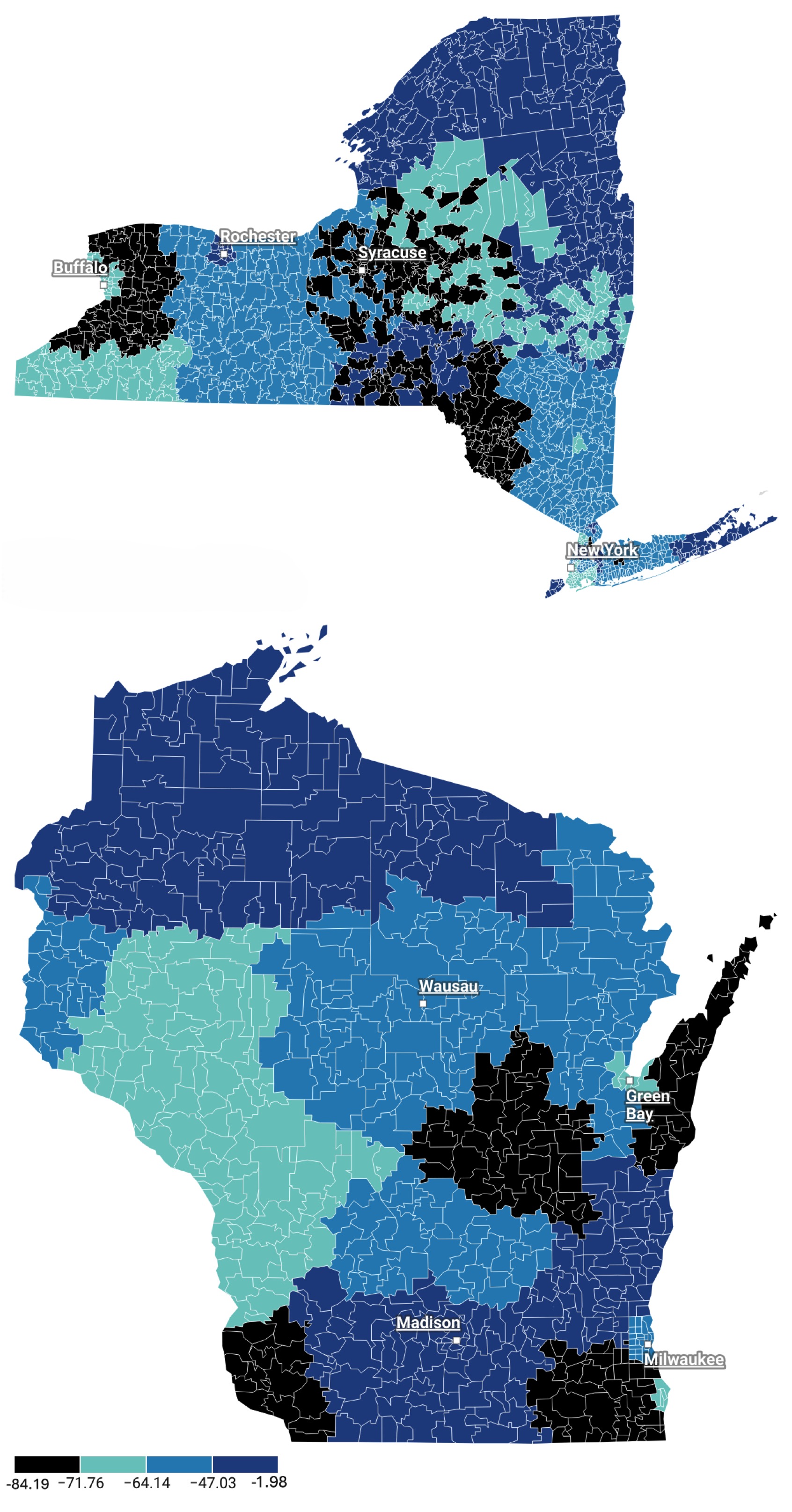
